# The use of data for planning and service improvement in Tanzanian Primary healthcare facilities: Experience from Star Rating Assessment

**DOI:** 10.1101/2022.06.03.22275952

**Authors:** Chrisogone C. German, Talhiya A. Yahya, Joseph C. Hokororo, Erick S. Kinyenje, Saumu I. Nungu, Mohamed A. Mohamed, Mbwana M. Degeh, Omary A. Nassoro, Syabo M. Mwaisengela, Radenta P. Bahegwa, Yohanes S. Msigwa, Ruth R. Ngowi, Laura E. Marandu, Eliudi S. Eliakimu

## Abstract

**Background:** The use of data for planning and improving healthcare delivery is sub-optimal among developing countries. In 2015, Tanzania started to implement Star Rating Assessment (SRA) process for primary health care (PHC) facilities to improve various dimensions of quality of services, including the use of data. We aimed at assessing the extent and predictors of data use in Tanzanian PHC facilities.

**Methodology:** We used the most current national SRA data available in DHIS2 that was collected in 2017/2018 from all 7,289 PHC facilities. A facility was considered using data if gained 80% of the allocated scores. Other dependent variables were the three components that together contribute to the use of data [If PHC facility has Health Management Information systems (HMIS) functional, disseminate information, and has proper medical records]. We determined the association between data use and facility ownership status (public or private), location of the facility (rural or urban) and facility service level (dispensary, health centre or hospital). Results are presented as proportions of facilities that qualified for data use and the three components. The associations are reported in Adjusted odds ratio (AOR) with a 95% confidence interval (CI).

**Results:** A total of 6,663(91.4%) PHC facilities met our inclusion criteria for analysis. Among the facilities: 1,198(18.0%) had used data for planning and services improvement; 3,792(56.9%) had functional HMIS; 1,752(26.3%) had disseminated data; and 631(9.5%) had proper medical records. PHC facilities that are publicly owned (AOR 1.25; 95% CI: 1.05–1.48) and those at higher service level [hospitals (AOR 1.77; 95% CI: 1.27–2.46) and health centres (AOR 1.39; 95% CI: 1.15–1.68) compared to dispensaries] were more likely to use data.

**Conclusion:** The *use of facility data for planning and services improvement* in Tanzanian PHC facilities is low, and much effort needs to be targeted at privately-owned and low-level PHC facilities.

## INTRODUCTION

Having a culture of data use at all levels of health services delivery is an essential element for health system strengthening [1]. The recent report of The Lancet Global Health Commission on high-quality health systems in the sustainable development goals era has noted that “*health systems need to develop the capacity to measure and use data to learn*” [2].

However, data use in health sector in low- and middle-income countries (LMICs) is sub-optimal. Issues affecting data use have been grouped into three areas – organizational, behavioural, and technical issues [3, 4]. In a study that documented a seven years experience of interventions in Mozambique, Rwanda, and Zambia, it was recommended that to improve data used for quality improvement; it is important to conduct data quality assessments and to improve the skills of service providers in data analysis and visualization [5]. Also, implementation of a combination of interventions that target behavioural and technical factors have been shown to improve data quality and use [6]. Involving local stakeholders in designing data collection, has been reported to be successful as measured by completion and accuracy of data collection tools [7].

In a systematic review that looked at challenges in the use of routine health information data in LMICs countries, Hoxha and colleagues identified organizational or environmental challenges as the most commonly reported barriers to data use [3]. Some interventions have been shown to have the potential for helping to establish a culture of data use including decision-makers/supervisors being examples to others (role models) through advocating for data use at the district/council level; using incentives such as the implementation of performance-based financing; and use of accountability system through the use of open performance review and appraisal system [8].

In Uganda, challenges found in health centres in implementing data quality assurance of routine information systems included: “*laborious and tedious manual system; the difficulty to archive and retrieve records; insufficient health management information system forms; and difficulty in delivering hard copies of reports to relevant stakeholders*” [9].

In 2015, Tanzania started to implement Star Rating Assessment (SRA) process for primary health care (PHC) facilities to improve the quality of services provided [10]. The assessment used a set of tools separate for dispensaries, health centres, and level one hospitals (hospitals at the council), with a total of 12 services areas in which service area three is on “*use of facility data for planning and services improvement*” [10, 11].

This study aimed to assess the extent of data use in PHC facilities in terms of: functionality of the health management information system (HMIS); information use and dissemination; and medical records status (recording and retrieval, as well as confidentiality of patient’s records).

## METHODS

### Conceptual framework

The design of the SRA Tools in service area 3 adapted the “*performance of routine information system management* (PRISM)” framework developed by Aqil and colleagues [12]. The elements assessed matched with the components of the PRISM framework are shown in table 1. The outcome measure which is improvement in data use for planning and services improvement is related to the outcome component of the PRISM framework which is “*improved Routine Health Information System* (RHIS)”. Therefore, based on the core components of the PRISM framework (technical, organizational and behavioural factors) matched with the assessed components of service areas 3 (Table 1), we conceptualized that improved data use for planning and improvement of services in primary health care facilities is a function of the functionality of HMIS, information use and dissemination, and medical records in a particular facility, and demographics of health facilities, as shown in Figure 1. The extent of the components of the PRISM framework covered in service area 3 of SRA Tools has the potential to provide comprehensive information that will inform the improvement of data used for planning and improvement of service delivery in PHC facilities in Tanzania [1].

**Table 1:** Data use areas assessed matched with the PRISM framework

| **3.0. Use of facility data for planning and services improvement** |  | **PRISM framework components** |
| --- | --- | --- |
| **Service area 3 components** | **Star Rating Indicators** |  |
| 3.1. Functionality of HMIS | Staff training on HMIS | Organizational factors |
|  | HMIS tools in use and filled correctly | ▪ Technical factors<br>▪ Organizational factors<br>▪ Behavioral factors |
| 3.2. Information dissemination | Data interpreted and used at the facility | Organizational factors<br>RHIS processes |
|  | Facility profile report shared with village and wards | Organizational factors |
| 3.3. Medical record | Recording and retrieval of medical records | Technical factors<br>RHIS processes |
|  | Confidentiality assured for patient records | RHIS processes<br>Organizational factors |

**Table 1.** Predictors associated with data use in primary healthcare facilities.

| Variable | Data use in the facility |  |  |  | Bivariate |  |  | Multivariate |  |  |
| --- | --- | --- | --- | --- | --- | --- | --- | --- | --- | --- |
|  | Yes | % | No | % | COR | 95% CI | p-value | AOR | 95% CI | p-value |
| **Facility type** |  |  |  |  |  |  |  |  |  |  |
| Health Centre | 160 | 21.9 | 572 | 78.1 | 1.34 | 1.11-1.62 | 0.002 | 1.39 | 1.15-1.68 | 0.001* |
| Hospital | 52 | 24.8 | 158 | 74.2 | 1.58 | 1.15-2.18 | 0.005 | 1.77 | 1.27-2.46 | 0.001* |
| Dispensary | 986 | 17.2 | 4,735 | 82.8 | Ref |  |  | Ref |  |  |
| **Ownership** |  |  |  |  |  |  |  |  |  |  |
| Public | 972 | 18.5 | 4,291 | 81.5 | 1.18 | 1.00-1.38 | 0.044 | 1.25 | 1.05-1.48 | 0.012* |
| Private | 226 | 16.1 | 1,174 | 83.9 | Ref |  |  | Ref |  |  |
| **Location** |  |  |  |  |  |  |  |  |  |  |
| Rural | 933 | 18.2 | 4,197 | 81.8 | 1.06 | 0.92-1.24 | 0.420 | 1.05 | 0.90-1.23 | 0.549 |

*p*- Values are calculated using chi-square test
\*Predictors whose association were found significant in the final logistic regression model
COR=Crude/unadjusted Odds Ratio, AOR=Adjusted Odds Ratio

**Figure 1:**
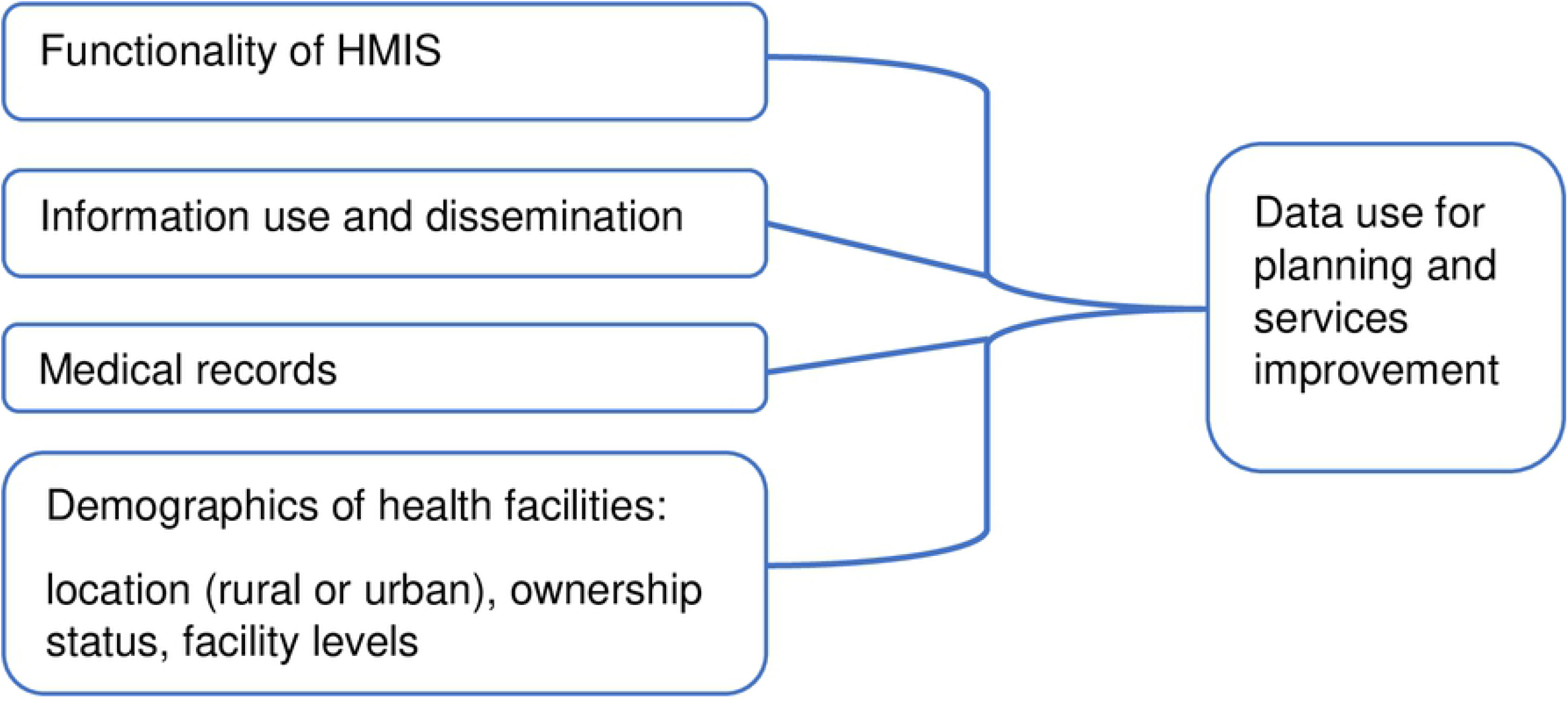
Conceptual Framework on data use for planning and service improvement in PHC facilities in Tanzania.

### Study Objectives

The objectives of our study were to determine the proportion of PHC facilities: that use data for planning and services improvement; that use HMIS tools correctly; that disseminate information; with proper medical records; and determine predictors associated with data use in PHC facilities.

### Study design

This was a retrospective cross-sectional study that used secondary data related to the use of facility data for planning and services improvement that are found in a national data warehouse known as District Health Information System 2 (DHIS2). This data was collected during the Star Rating assessment that was conducted in 2017/2018.

### Target population

All operating PHC Facilities in Tanzania (Dispensaries, Health Centers and Hospitals at Council level) regardless of their ownership, i.e., public (Local Government Authority [LGA], Military, Police, Prisons, Parastatals and other Ministries, Departments and Agencies [MDAs]) or private (Faith-Based Organization [FBO], Non-Governmental Organization [NGO], Private for Profit).

### Study population

All PHC facilities which were assessed during reassessment held in 2017/2018. The assessment was conducted in all 7,289 PHC facilities by then from all 26 regions of Tanzania Mainland. PHC facilities constitute the majority of healthcare facilities (more than 97%) in Tanzania.

#### Inclusion criteria

All facilities that were assessed in 2017/18 and whose performance and characteristics were able to be identified from the DHIS2 after data cleaning.

#### Exclusion criteria

Facilities with incomplete data were excluded during the analysis.

### Data sources and collection

#### Data collection

This section explains how data that make up the SRA dataset in the DHIS2 were collected. Data collection from each facility was done by at least four trained personnel; each from all healthcare administrative levels, i.e., national, regional, council and facility-level to ensure transparency and fairness. The assessors captured the score per question at the facility level using the SRA Tool (SRT) and subsequently entered the values in Microsoft Excel Sheets to produce the score per each service area, including the area under this study, i.e., the use of facility data for planning and services improvement.

As presented in Table 2, the section on the use of facility data for planning and improvement in SRT is grouped into three components each made of two indicators. Table 2 presents questions and assessment criteria that were used to score the above variables during SRA. For each question, one response was chosen among the following possible scoring options; Yes’ (score=1), Partial (score=0.5) or ‘No’ (score=0).

**Table 2:**
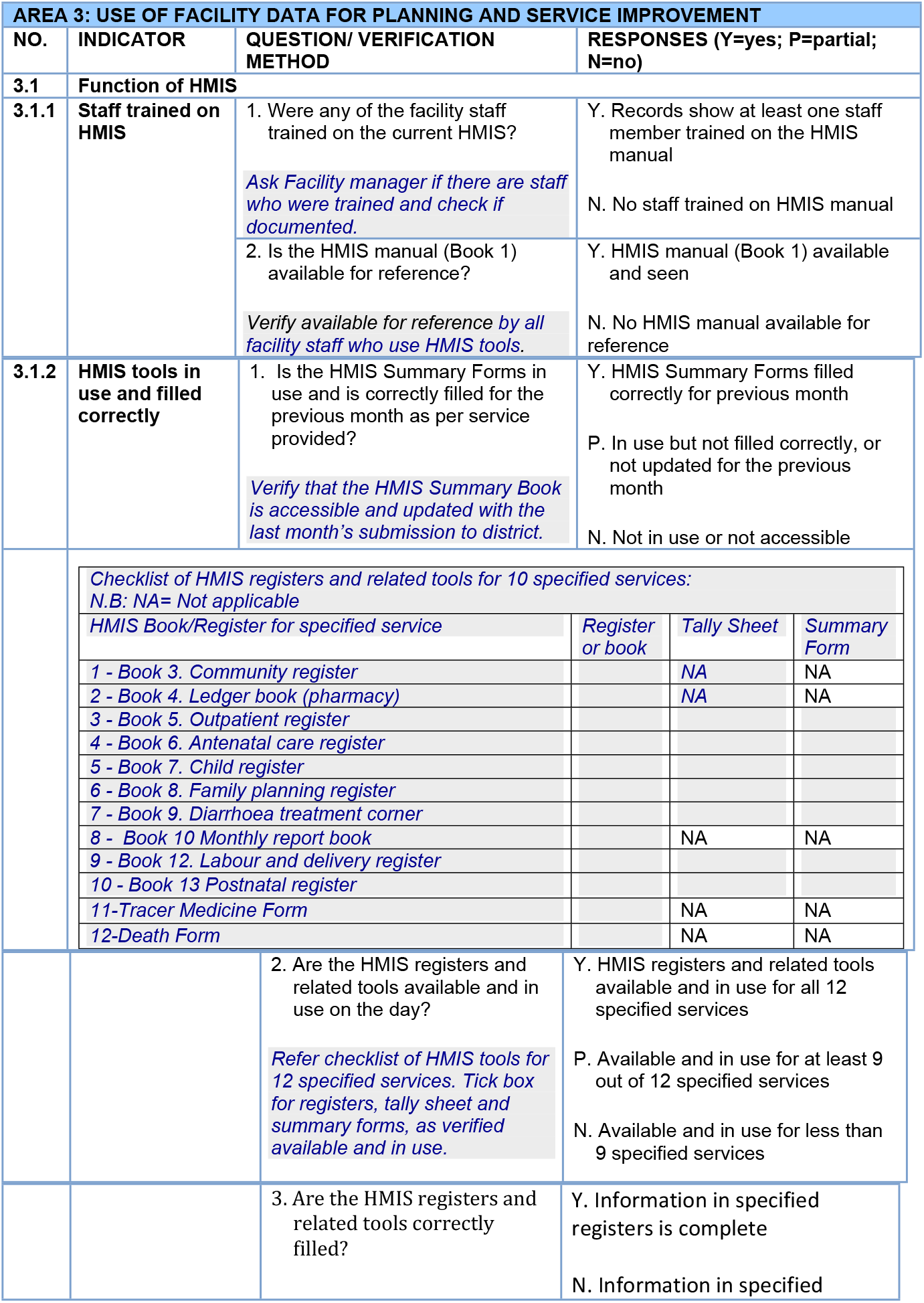
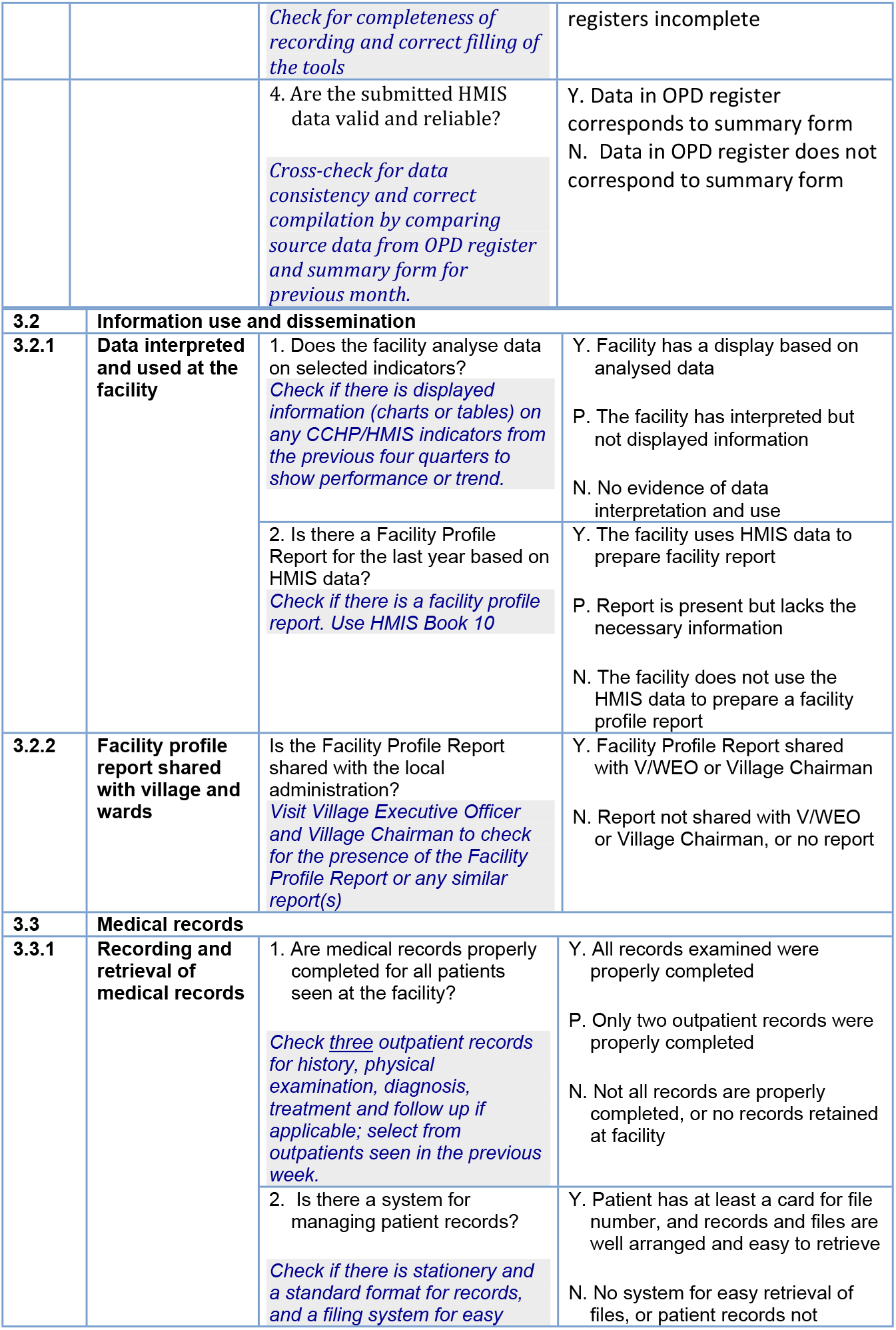
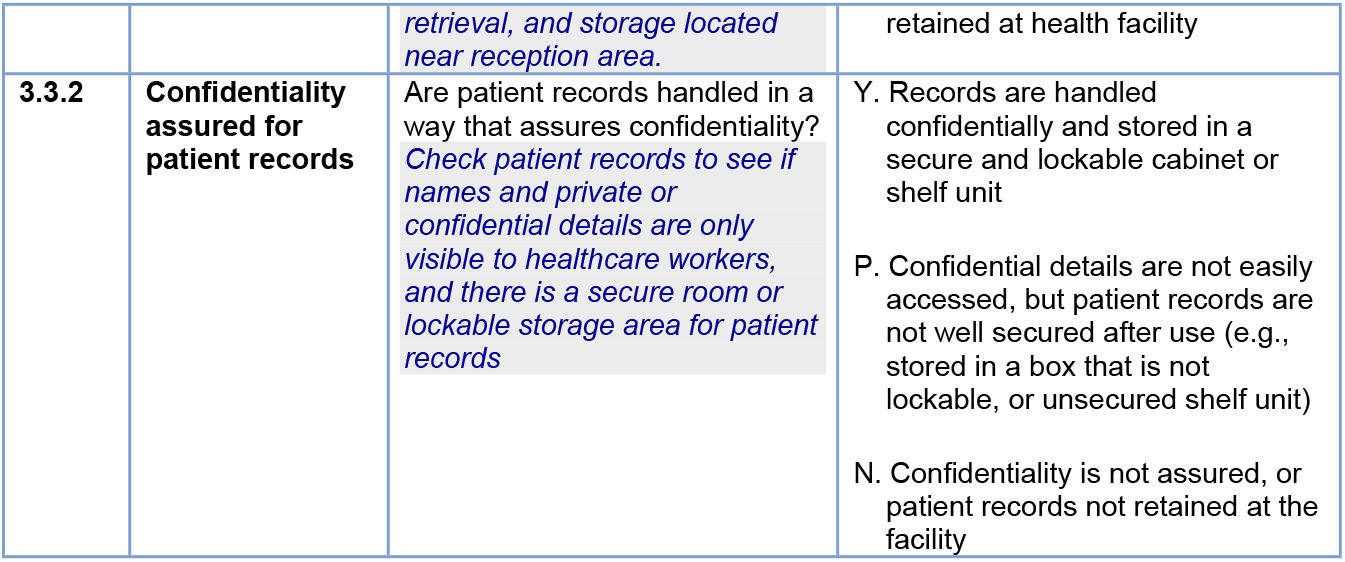
Specific questions asked for each indicator and the assessment criteria

Data from the sheets were cleaned and compiled at district level immediately to ensure correctness, completeness, accuracy and timeliness of the data. The SRA at national level coordinated by Health Services Inspectorate and Quality Assurance Section of the then Health Quality Assurance Division, which is currently Health Quality Assurance Unit (HQAU) of the Ministry of Health [13].

#### Data sources

The targeted data were extracted from the DHIS2 database in form of Microsoft Excel Sheets. The sheets were checked for data quality and cleaned before use.

### Study variables

The main dependent variable of interest for this study was the use of facility data for planning and services improvement and was used to determine its association with independent variables, i.e., the facility-related variables, i.e., facility ownership status (public or private owned), location of the facility (rural or urban-based) and health facility service level (dispensary, health centre or hospital level one). Other dependent variables were the three components that contribute to data use that are mentioned in Table 1, i.e., functionality of HMIS, data dissemination and medical records.

### Data management and analysis

The facilities were characterized based on ownership status, location and health service level. Data cleaning and analysis was done using STATA 15. The data was cleaned and checked for completeness and outliers before analysis. The performance in each dependent variable was measured as the proportion of PHC facilities that scored at least 80% from allocated points. This cut-off point is provided in the National Guidelines for Recognition of Implementation Status of Quality Improvement Initiatives in Health Facilities [14].

Therefore, a facility could gain 12 points maximum from 12 questions and they needed 9.6 to be qualified as using data for planning and improvement. Likewise, a facility needed 4.6 points out of 6 points to have a functional HMIS. Also, 2.4 points were needed for both data dissemination and medical recods variables for the facility to qualify because these variables are made of three assessment questions each.

We determined an association between the binary variable and independent variables (i.e., facility’s ownership, facility service level and location) to determine the predictors of data use for planning and improvement at the PHC facilities. The association was measured by calculating the odds ratio with a 95% confidence interval and a *P*-value of < 0.05 was considered as statistically significant.

## RESULTS

A total of 6,663 PHC facilities; which is equivalent to 91.4% of all 7,289 PHC facilities that were included in the SRA assessment of 2017/18 [10, 11] met inclusion criteria and were included in the final analysis under this study. Among the facilities; 1,198 (18.0%) had used data for planning and services improvement. These are the facilities that gained at least 80% in all assessment questions. Regarding the three components of data use for planning and improvement; 3,792 (56.9%) of the facilities had functional HIMS; 1,752(26.3%) had disseminated data; and 631(9.5%) had proper medical records.

As presented in Table 3; all facility-related variables that were included in our study were significantly associated to “data use for planning and improvement at PHC facilities” during both bivariate and multivariate analysis except the location of the facility. In comparison to the lowest level of PHC facilities, i.e., dispensary-level; findings have shown the higher the level of PHC facilities the higher likelihood of using data for planning and improvement [i.e., health centres (AOR 1.39; 95% CI: 1.15–1.68), hospital-level (AOR 1.77; 95% CI: 1.27– 2.46)]. Public-owned PHC facilities had a 25% increased likelihood of using data for planning and improvement compared to private-owned facilities (AOR 1.25; 95% CI: 1.05–1.48).

## DISCUSSION

The findings have shown health facility data are used for planning and improvement in only one-fifth of the PHC facilities in Tanzania. Among three components that determine the use of data in the PHC facilities; most of facilities failed in having proper medical records whereby only one-tenth of the facilities had capability to record, retrieve and ensure patient records are handled in a way that assures confidentiality. More than half of the facilities had HMIS system functional and therefore this was relatively the most performed component among the components that determine data use in the health facilities. The national target is at least 80% and therefore effort is still needed to improve its functionality. The facility had HMIS functional if staff are trained in HMIS and HMIS tools are used and filled correctly. Moreover, about a quarter of the facilities had analysed data and displayed it for public view. A study in five countries (Cambodia, Ghana, Mozambique, Nigeria and Tanzania) has documented the burden of recording and reporting in PHC facilities. The findings show that the functionality of HMIS and medical records in PHC facilities in Tanzania may be affected by the burden of recording and reporting due to the volume of registers in which for completing monthly reporting forms it takes up to 65-hours, with 29 reporting forms [15]. In the light of the above challenges, data usage in Tanzania has been further emphasized in publicly-owned PHCs through Direct Health Facility Financing (DHFF) initiatives that were targeting public facilities since 2017 [16]. Among others, DHFF requires public facilities to collect quality data that will be used for planning interventions that will target specific local needs. This may be the reason why public facilities have performed better in this study compared to private ones.

In a study which was conducted in 12 health facilities in three Tanzanian regions (Arusha, Lindi, and Geita) to assess the capacity of health workers to analyze and use data for family planning services, they found that: health workers have inadequate skills for data analysis and computer use; the facilities have a weak culture of data analysis and use; lower-level health facilities, lacked internet access, hence affecting their access to DHIS2; and lack of data ownership among health workers thinking in which they believed that data generated at health facilities belong to the Council Health Management Team (CHMT) and not the facilities; and lack of training on collecting, analyzing, presenting, and using data [17]. Also, in a study conducted in 11 districts involving 115 health facilities in Tanzania, it was found that poor data use was common in most of the districts, due to inadequate data management skills, few facilities receiving supervision visits and feedback from CHMT, and lack of training [18].

A study in Nigeria found that training of health workers in PHC facilities improved their data management skills including filling registers and forms, data analysis and use [19]. Findings from a study in Ethiopia have shown slightly more than half of 250 staff who participated in the study reported to use facility data routinely to develop plan; while identified factors that hindered data use included residence, data management knowledge, work load, computer skill, computer access, supportive supervision, HMIS training, and availability of HMIS guideline and formats [20]. Also, research evidence in Tanzania shows that the level of education is positively associated with the use of HMIS especially if the systems are electronic [21] and therefore, we presume the use of data in high-level facilities was due to the presence of relatively more educated staff in these facilities. In Costa Rica, a strong culture of valuing data as a tool to drive improvements coupled with technical and managerial support has contributed to the use of data for improving care to patients and improving population health [22]. Rendell and colleagues (2020) have identified several factors that can influence data use in PHC services which they organized them into three groups: *governance (leadership, participatory monitoring, regular review of data); production of information (presentation of findings, data quality, qualitative data); and health information system resources (electronic health management information systems, organizational structure, training) [23]*.

The ultimate goal of SRA processes is to improve performance management of the PHC facilities that includes management of health information systems and use of data [10]. Therefore, the government of Tanzania must strongly uphold and implement the digital health strategy namely “The National Digital Health Strategy 2019–2024”-which is also in line with both the Tanzania Development Vision 2025 and the Health Sector Strategic Plan 2021–2026 [24, 25]. The strategy aims at improving the management of health information systems in the PHC facilities particularly through the use of sustainable and interoperable electronic systems. Improving the use of data is essential in strengthening PHC in a way that will ensure its orientation towards person centred care in PHC [26]. Therefore, it is recommended for Tanzania to make use of the data use partnership project to strengthen culture of data use in PHC facilities by focusing on having better data (i.e., strengthening quality of data production) and implementing interventions that will help to nudge health workers towards a behaviour of regular data use [27]. By strengthening the use of data in PHC facilities, it will be an important step in building a learning health system in Tanzania [28, 29].

## CONCLUSION

The use of facility data for planning and services improvement in PHC facilities is low in Tanzania and as result the quality of care could be hampered. Much efforts need to be targeted at privately-owned and low-level PHC facilities (i.e., dispensaries). The facilities performed more or less equal despite locality and therefore more research is needed to explore other factors that could influence the use of data in PHC facilities in Tanzania. The very low level of PHC facilities that have proper medical records require special attention in order to ensure that there is proper recording, retrieval and privacy of patients’ records.

## Data Availability

All relevant data are within the manuscript and its Supporting Information files.

## Acknowledgement

The authors are passing their sincere gratitude to the Ministry of Health especially, the Health Quality Assurance Unit for permitting us to use the SRA data.

Apart from government institutions, the authors extend appreciation to development partners such as World Bank, The United States - Centres for Disease Control and Prevention (CDC), Danish International Development Agency (DANIDA), and The World Health Organization whom together made SRA possible. Others were the communities of the facilities visited, PharmAccess International, Association of Private Health Facilities in Tanzania (APHTA), Christian Social Services Commission (CSSC), and Development Partners in Health-Group (DPG-H).

